# External quality assessment of BCR-ABL1 quantification in chronic myeloid leukemia: a pilot interlaboratory study from Russia

**DOI:** 10.1101/2025.11.14.25340242

**Authors:** Elena V. Khoroshun, Kuvat T. Momynaliev, Igor V. Ivanov

## Abstract

BCR-ABL1 is a chimeric oncogene underlying chronic myeloid leukemia (CML) and serves as a key marker for monitoring minimal residual disease. Quantitative RT-qPCR analysis of BCR-ABL1 transcripts enables assessment of molecular response depth and timely detection of treatment failure or resistance. In international practice, the introduction of the International Scale (IS), along with method standardization, has significantly improved the comparability of results between laboratories. Participation in external quality assessment (EQA) programs is a mandatory accreditation requirement in countries with established CML monitoring systems, allowing laboratories to evaluate analytical accuracy and to adjust protocols when deviations are detected. Until recently, Russia lacked both standardized IS calibration and a national EQA program for BCR-ABL1 testing, which hindered the assessment of interlaboratory precision. This pilot study represents the first interlaboratory comparison of quantitative BCR-ABL1 measurement in the Russian Federation. Two panels of lyophilized control samples (five levels each, from M1 to M4.5) were distributed and analyzed by 14 laboratories (15 datasets) using their routine protocols without IS conversion. Interlaboratory variability was high: the overall coefficient of variation ranged from approximately 40 percent to 50 percent, with individual laboratory results at the high level (about 10 percent) varying from 3 percent to 27 percent. Most laboratories’ results were within plus/minus 3 standard deviations of the consensus median, but two laboratories showed consistent overestimation (bias about +3.8 percentage points) or underestimation (bias about −1.7 percentage points), leading to multi-fold errors at level M1. These findings highlight the urgent need to implement IS calibration and to establish a national EQA program for BCR-ABL1 testing in Russia to improve result comparability. Adoption of these measures will help align molecular monitoring of CML in Russia with international standards and strengthen clinician confidence in laboratory testing

## Main text

Quantitative measurement of BCR-ABL1 transcripts by RT-qPCR is the cornerstone of molecular response monitoring in CML [1–3]. In international practice, the adoption of a unified International Scale (IS) along with standardized methods has significantly improved the comparability of BCR-ABL1 results across laboratories [4–6]. Furthermore, regular participation in external quality assessment (EQA) programs (e.g., CAP in the US, UK NEQAS in the UK) is mandatory for laboratory accreditation in countries with established CML monitoring [7–11]. Such programs allow each laboratory to verify its measurement accuracy and prompt those with outlying results to adjust their methods and calibration.

Until recently, the Russian Federation had no unified system for converting BCR-ABL1 results to the IS and no national EQA program for BCR-ABL1 testing. This lack of standardization has made it difficult to evaluate interlaboratory accuracy and compare CML monitoring data within the country. We report the first attempt at an interlaboratory comparison of BCR-ABL1 quantitative testing in Russia. The goal was to assess variability in results between laboratories, identify problem areas, and substantiate the need to implement the international scale and to participate in external quality control programs.

For this study, two panels of lyophilized control samples (designated panel 1 and panel 2) were prepared, each comprising 5 samples with different BCR-ABL1 transcript levels corresponding to molecular response (MR) levels M1 through M4.5. Each panel thus contained samples at approximately the following %BCR-ABL1/ABL1 ratios: ~10% (M1), ~1% (M2), ~0.1% (M3), ~0.01% (M4), and ~0.003–0.005% (M4.5), covering the range from no significant molecular response to deep MR4.5. These samples were produced by mixing cells from K562 (BCR-ABL1 e14a2-positive) and HL60 (BCR-ABL1-negative) cell lines in varying proportions, followed by lyophilization for stability. This approach mirrors the design of EQA panels used in other countries. Each sample was labeled OK-M1, OK-M2, … OK-M4.5.

A total of 14 laboratories from various regions of Russia participated, providing 15 data sets (one lab submitted results from two independent methods, counted separately). Each lab extracted RNA from the provided samples and performed RT-qPCR according to its routine protocol. A variety of amplification platforms (e.g., Rotor-Gene, CFX96, DTprime) and reagent kits commonly used in Russia (such as the domestic “AmpliSens Leucosis Quant” kit) or equivalent in-house methods were employed. None of the laboratories had the capability to convert their results to %IS due to the lack of conversion factors or certified IS reference standards in Russia at the time. Therefore, all results were reported simply as the percentage of BCR-ABL1 relative to a control gene (typically ABL1) without IS transformation.

Each laboratory measured each sample in duplicate (two replicates per sample in panel 1 and similarly in panel 2). For analysis, the mean %BCR-ABL1/ABL1 value of replicates was calculated for each lab on each sample (if one replicate was missing, the single available value was used). Using the aggregate of all labs’ results, we determined assigned consensus values for each control level M1–M4.5 by taking the median of all laboratory results for that sample. The median was chosen as a robust estimate of the true value in the absence of an established calibrator. We then assessed each lab’s deviation from the consensus by computing z-scores: *z* = (*x*_lab_ - median)/*σ*, where σ is a robust standard deviation estimate (the median absolute deviation, MAD) for that sample’s results. Results with |z| > 3 were considered significant outliers (beyond acceptable limits). In addition, to quantify each laboratory’s overall accuracy, we calculated the bias (mean difference between the lab’s results and the assigned values across all samples), the mean absolute error (MAE; average of absolute differences), and the root mean square error (RMSE) for each lab. These metrics, expressed in the same units (percentage points of BCR-ABL1), indicate each lab’s systematic error (positive or negative bias), typical error magnitude (MAE), and sensitivity to large errors (RMSE). Data visualization included boxplots to show result distributions at each control level and a heatmap of z-scores (equivalently, relative biases) for each lab across all levels.

## Results

The assigned consensus values and overall variability are summarized first. Combining results from both panels, the median %BCR-ABL1 values for levels M1, M2, M3, M4, and M4.5 were approximately 10.1%, 1.02%, 0.136%, 0.010%, and 0.005%, respectively. As expected, these values closely correspond to the target MR levels (for example, M3 ~0.1% ≈ major molecular response, MMR). However, the spread of individual laboratory results was considerable. For the highest level M1 (~10%), the participant labs’ mean values ranged from ~3% up to ~27%. At the M2 level (~1%), lab means spanned ~0.19% to ~2.12%, and at M3 (~0.1%) from ~0.018% to ~0.235%. Even at the very low M4–M4.5 levels, appreciable variation persisted – for example, for a true ~0.005% sample, reported values ranged ~0.003–0.011% (**Figure 1**). The interlaboratory coefficient of variation (%CV) was on the order of 40–47% across the control points, reflecting the lack of a uniform calibration or method. By comparison, in the first Chinese multi-center EQA for BCR-ABL1 (66 labs), an even larger variability was noted (CV up to 60–100% without IS calibration), with a trend toward reduction (to ~62% CV) among labs that converted results to the IS [8].

**Fig. 1.**
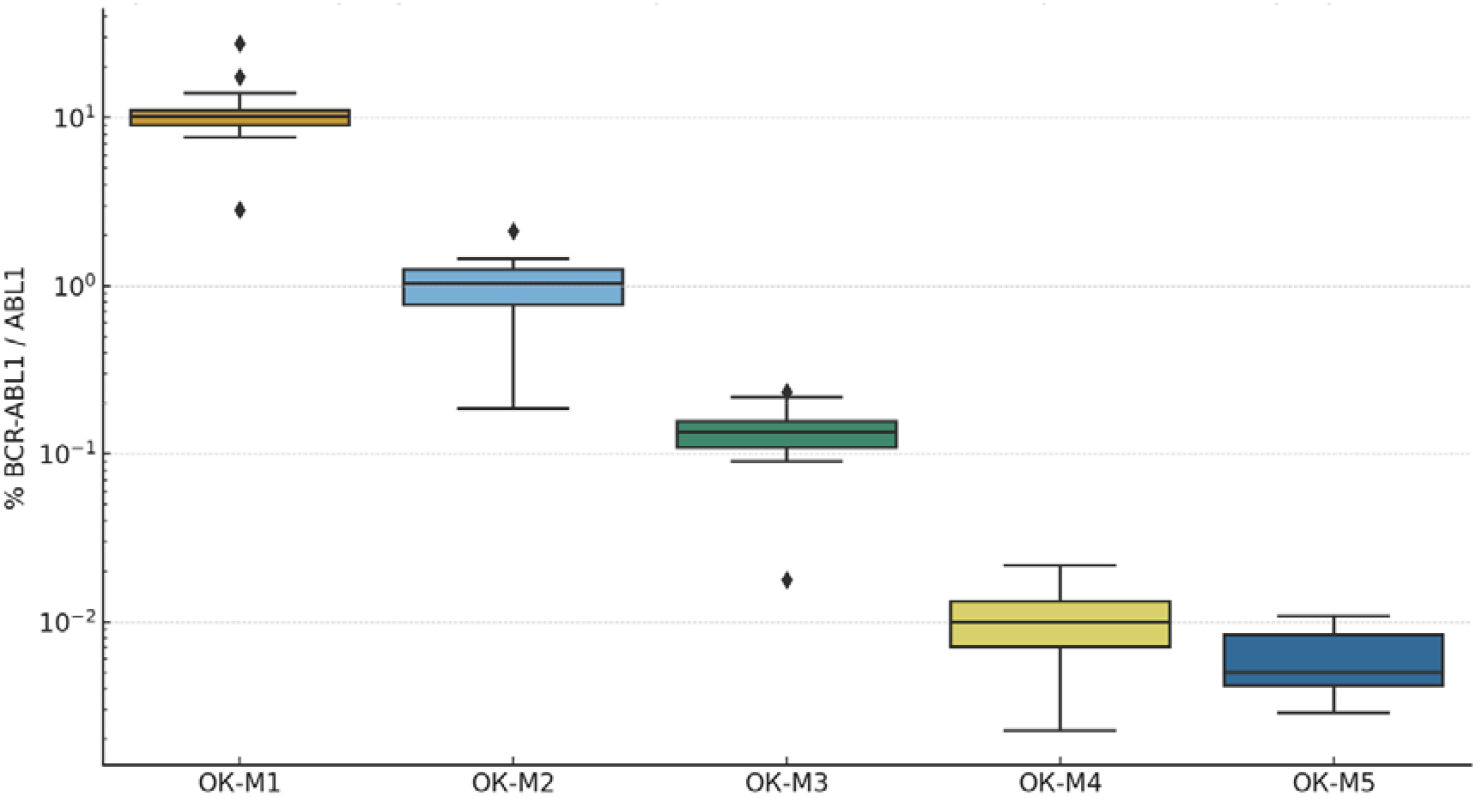
Distribution of BCR-ABL1 measurement results (% relative to control gene) for five control levels (M1–M4.5) across all participating laboratories. Boxes show the interquartile range and median; whiskers show the range excluding outliers. *Y*-axis is on a logarithmic scale. As BCR-ABL1 concentration decreases (from M1 to M4.5), the absolute interlaboratory spread in results diminishes, but the relative variability remains high due to proximity to the detection limit. The greatest absolute spread is observed at level M1 (~10%): the interquartile range is ~9–11%, and individual laboratories reported values outside these bounds.

Next, we evaluated individual laboratory biases and identified specific issues (**Figure 2**). Most labs showed acceptable agreement with the consensus (|z| ≤ 2–3 at all levels). However, two laboratories exhibited clear systematic errors. One lab (code #8) consistently overestimated BCR-ABL1 at all levels, with an average bias of +3.75 percentage points (pp) – meaning this lab’s results were on average 3.75 pp higher than the true values. At the ~10% level (M1), lab 8 reported ~27% BCR-ABL1 instead of ~10%, an ~2.7-fold high bias (z ≈ +11.6), which appears as a bright red outlier on the heatmap. Another lab (code #12) conversely underestimated results, with mean bias –1.65 pp (i.e. a systematic low bias. At M1, lab 12 reported ~2.8% instead of ~10% (~3.5-fold low, z ≈ –4.8, a bright blue block on the heatmap). Thus, at least two labs made fundamental systematic errors: in the first case likely an issue of calibration or contamination causing over-quantification, and in the second perhaps poor RNA extraction or amplification efficiency, or a calculation error leading to under-quantification. Aside from these, one lab (code #3) produced an outlying result at M1 (z ≈ +4.9), though its other levels were closer to consensus; this suggests a one-time technical error (e.g. pipetting) with the highest-concentration sample. At the low transcript levels (M4–M4.5), several labs showed within-lab inconsistency: for example, lab #13 obtained highly divergent replicates for the M4.5 sample (0.021% vs 0.0037% BCR-ABL1), yielding z-scores from +2 to –0.3 (seen as alternating red and blue in row 13 for levels M4–M4.5). Lab #14 had a similar reproducibility problem at the highest and lowest levels (its two replicate results differed greatly, though the average still fell near the median; on the heatmap, lab 14 does not show a strong color, indicating the error canceled out upon averaging). These cases highlight the importance of internal reproducibility control, especially near the detection limits of the assay.

**Fig. 2.**
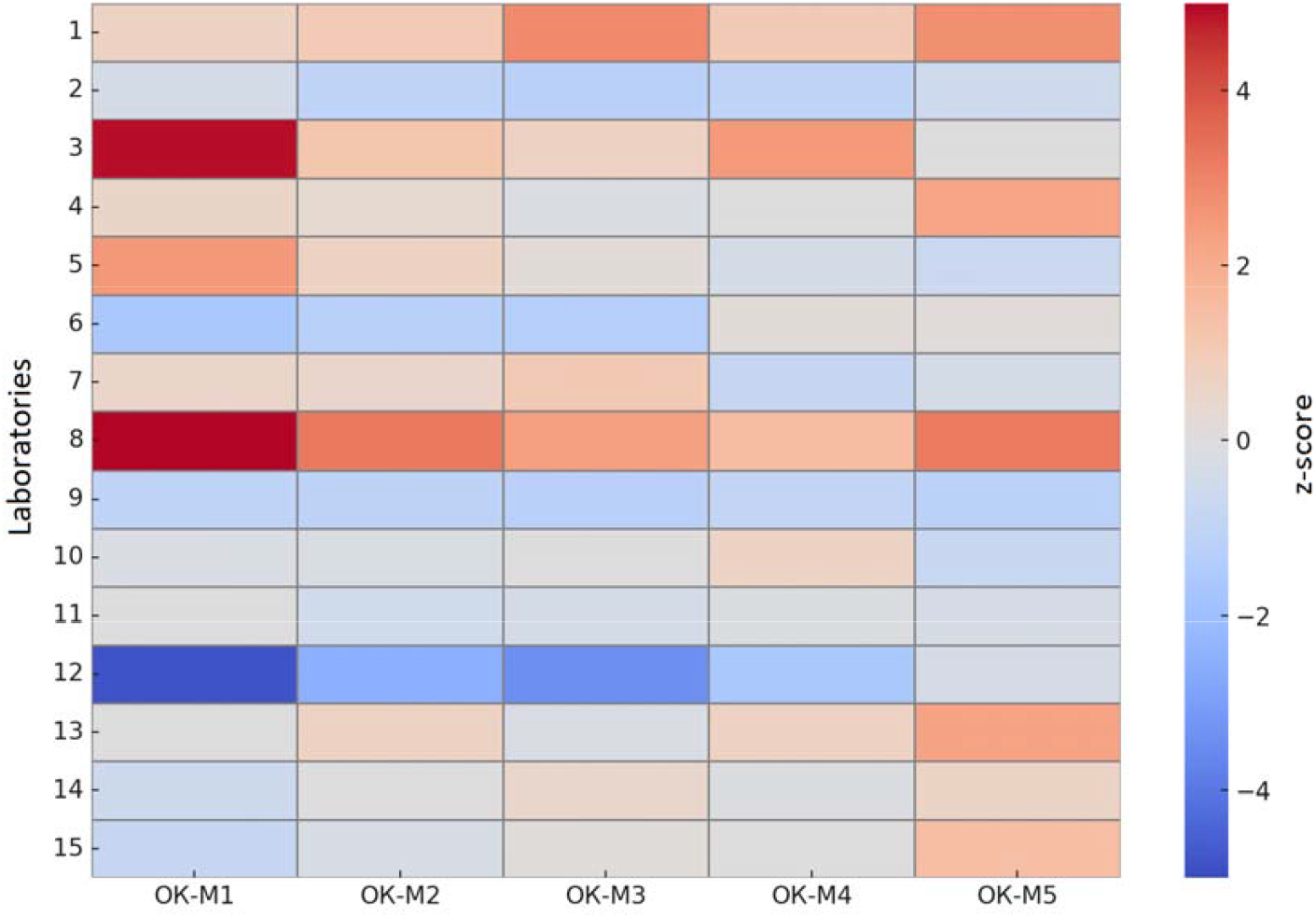
Heatmap of z-scores for the results of 15 laboratories across five control samples (M1– M4.5). White indicates exact agreement with the median (zero deviation); red shades indicate results higher than the consensus (positive bias), blue shades indicate lower results (negative bias). Color intensity is proportional to the magnitude of deviation (z-score scale shown on the right); the black dashed line marks the ±3σ limits. Most laboratories’ deviations fall within ±2–3σ (gray background), with the exception of two systematically biased profiles – bright red (lab #8) and bright blue (lab #12).

To provide an overall quantitative assessment of each laboratory’s accuracy, we computed the summary metrics bias, MAE, and RMSE across all 10 sample measurements (5 levels × 2 panels) per lab (**Table 1**). Most laboratories showed only small systematic bias (|bias| < 0.3 pp) and a mean absolute error on the order of 0.2–0.8 pp (which corresponds to roughly 10–20% relative error at the M1 level). The best performance was observed in labs #10 and #11 – their bias was near zero and their errors were minimal (MAE and RMSE ~0.07–0.12 pp), indicating results virtually coincident with the consensus values. In contrast, labs #8 and #12 stand out with much larger errors: lab 8 had MAE = 3.75 pp and RMSE = 7.86 pp (several-fold higher than others due to its systematic overestimation), and lab 12 had MAE = 1.65 pp, RMSE = 3.27 pp. Lab #3 also showed a notable average deviation (bias ~+1.57 pp, MAE 1.57 pp), attributable to its one large outlier at M1. These metrics align with the visual analysis in Fig. 2, confirming the presence of substantial interlaboratory discrepancies in the absence of a common standard.

**Table 1.** Summary accuracy metrics of BCR-ABL1 measurements by laboratory. *Bias* – mean deviation of the laboratory’s result from the assigned value (positive bias indicates a systematic high deviation, negative bias a low deviation); *MAE* – mean absolute error; *RMSE* – root mean square error. All values are in percentage points (% BCR-ABL1).

| Laboratory | Bias (pp) | MAE (pp) | RMSE (pp) |
| --- | --- | --- | --- |
| Lab 1 | +0.30 | 0.30 | 0.48 |
| Lab 2 | −0.19 | 0.19 | 0.30 |
| Lab 3 | +1.57 | 1.57 | 3.30 |
| Lab 4 | +0.20 | 0.20 | 0.40 |
| Lab 5 | +0.80 | 0.80 | 1.69 |
| Lab 6 | −0.58 | 0.58 | 1.10 |
| Lab 7 | +0.19 | 0.19 | 0.37 |
| Lab 8 | +3.75 | 3.75 | 7.86 |
| Lab 9 | −0.40 | 0.40 | 0.72 |
| Lab 10 | −0.07 | 0.07 | 0.12 |
| Lab 11 | −0.03 | 0.04 | 0.07 |
| Lab 12 | −1.65 | 1.65 | 3.27 |
| Lab 13 | +0.05 | 0.05 | 0.10 |
| Lab 14 | −0.18 | 0.18 | 0.40 |
| Lab 15 | −0.26 | 0.26 | 0.55 |

Accuracy was further analyzed based on the kits used. Eleven result sets were obtained using the commercial kit “AmpliSens Leucosis Quant,” while four sets were based on non-standardized (in-house) laboratory-developed protocols. Median bias values in both groups were close to zero, and the median MAE ranged from approximately 0.2 to 0.3 percentage points, indicating comparable accuracy across most laboratories. However, mean values revealed differences: the AmpliSens group showed higher bias and error, largely due to several laboratories with substantial deviations. This underscores the importance of using median-based metrics for interlaboratory comparisons, particularly in the presence of outliers. Thus, employing a standardized reagent kit does not automatically guarantee higher accuracy—the reliability of results depends on the correct implementation of the methodology in each individual laboratory.

The results demonstrate substantial interlaboratory variability in BCR-ABL1 quantification under conditions lacking standardized calibration. Similar challenges have been reported internationally: prior to the introduction of a unified International Scale (IS), discrepancies between laboratories often spanned several-fold, whereas IS conversion markedly reduced result dispersion [7–8]. Our findings confirm that the absence of IS calibration in Russian practice leads to similar large-scale differences between laboratories. Clearly, the lack of a common reference point (IS) contributed to the observed inconsistencies, and IS alignment is expected to significantly improve interlaboratory agreement. A limitation of this study is the relatively small number of participating laboratories (n = 14), as well as the lack of an absolute reference standard when assigning consensus values. Nevertheless, even under these conditions, significant deviations were observed in several centers. Expanding the program to include more laboratories and using certified reference materials (such as WHO international panels for %IS) would enable direct linkage of results to the IS [5–6].

In summary, this pilot study showed that the absence of standardization leads to a ~40–50% spread in results between laboratories. Some participants systematically over- or underestimated transcript levels by several-fold, which could potentially lead to incorrect clinical assessment of molecular response. Our data strongly support the urgent need to implement IS calibration and to establish a regular external quality assessment program for BCR-ABL1 testing in Russia. Implementing these measures would improve result comparability between centers, bring domestic CML monitoring practices in line with international standards, and enhance clinicians’ confidence in molecular test results.

## Data Availability

All data produced in the present study are available upon reasonable request to the authors

